# Knowledge and behaviors toward COVID-19 among U.S. residents during the early days of the pandemic

**DOI:** 10.1101/2020.03.31.20048967

**Authors:** John M Clements

## Abstract

**Objective:** To test the hypothesis that knowledge of COVID-19 influences participation in different behaviors including self-reports of purchasing more goods than usual, attending large gatherings, and using medical masks.

**Methods:** Cross-sectional online survey of 1,034 U.S. residents age 18+ conducted on March 17, 2020.

**Results:** For every point increase in knowledge, the odds of participation in purchasing more goods (OR=0.88, 95% CI:0.81-0.95), attending large gatherings (OR=0.87, 95%CI: 0.81-0.93), and using medical masks (OR=0.56, 95% CI:0.50-0.62) decreased by 12%, 13%, and 44%, respectively. Gen X and Millennial participants had 56% to 76% higher odds, respectively, of increased purchasing behavior, compared to Baby Boomers. Results suggest politicization of response recommendations. Democrats had 30% lower odds of attending large gatherings (OR=0.70, 95% CI:0.50-0.97), and 48% lower odds of using medical masks (OR=0.52, 95% CI:0.34-0.78), compared to Republicans.

**Conclusions:** This survey is one of the first attempts to study determinants of knowledge and behaviors in response to the COVID-19 pandemic in the U.S. A national, coordinated effort at pandemic response may ensure better compliance with behavioral recommendations to address this public health emergency.

## Introduction

Some of the most important problems in the world require an understanding and acceptance of science by the general public, including addressing health problems such as the emergence of the novel coronavirus (SARS-Cov-2) and subsequent disease (COVID-19) transmission. SARS-Cov-2 first emerged in December 2019 in Hubei Province, near Wuhan, China^1^. By mid-January 2020, Thailand and Japan were the first countries outside of China to report COVID-19 cases^1^. The Chinese government subsequently quarantined the greater Wuhan area on January 23, 2020 to prevent COVID-19 spread^2^.

On January 21, 2020, the first COVID-19 case in the United States was reported in Washington State^3^ and it was later reported that public health officials thought the virus was prevalent in the community for at least several weeks prior^4^. In the United States, the federal government ordered certain flights from China halted, and screening of passengers from other locations at different ports of arrival^5^. The Centers for Disease Control and Prevention (CDC), and the National Institutes of Health (NIH), began making recommendations, based on the scientific knowledge of the situation, to limit social contacts, encourage wise use of medical supplies including masks, and assure the public about the reliability of the food and consumable goods supplies^6^. However, even after these recommendations, there were reports of college students waiting in long lines at bars to celebrate their campuses closing,^7^ people buying medical grade masks,^8^ and hoarding everything from toilet paper to eggs and milk,^9^ even as the President sought to reassure the public that the supply of food and supplies was secure^10^.

Scholarship on public understanding of science (PUS) aims to explain public understanding of, involvement in, and trust in science. In the face of the current pandemic, this requires the public to understand and trust those who are making recommendations to limit exposure and spread of illness. The deficit model of PUS posits that a lack of support for science (and a subsequent rejection of recommendations) is due to a lack of understanding about science, and if scientists can find a way to fill this knowledge deficit, then support for science will increase. A more contemporary view of PUS is that the public’s knowledge is not deficient, but rather there is a deficit in trust in science, and in scientific experts specifically. Because of an increasing lack of trust in these institutions, Solomon^11^ observed that there is an increased personal rejection of science which then leads to lower levels of scientific literacy and understanding of science. Low literacy and understanding may influence people to not follow recommendations for addressing science-based problems, as evident with the current pandemic.

Much of the PUS literature examines trends in scientific knowledge (albeit self-reported knowledge, for the most part) and attitudes about science. Results are mixed as to whether increased knowledge leads to positive attitudes (variously described as trust, support, confidence, and support for funding) about science. Allum et al^12^ observed a small positive correlation between knowledge about science and positive attitudes about science and Miller^13^ reports that there is public support for science even in the face of a scientific literacy rate of 20%. The public’s support for science is necessary when addressing many important social issues, including an immediate need for the public to understand, and trust, the science about the novel Coronavirus pandemic currently plaguing the world. If the public does not trust the underlying science about these issues and does not trust in institutions that are tasked with managing this threat, it will be difficult to count on public support for policies to address these issues.

This paper describes a cross-sectional online survey designed to gauge public knowledge and behaviors about COVID-19 in the United States. Zhong et al^14^ conducted a similar study in China, approximately one week after the Hubei Province was put on lockdown (approximately eight weeks after the first case emerged), to determine the level of knowledge and public sentiment about the emerging pandemic in China. This study essentially replicates questions about knowledge from that study, while asking about more specific behvaiors, with a sample drawn from an online work platform (Amazon’s Mechanical Turk) to determine level of knowledge about COVID-19 and characteristics that influence knowledge and behaviors toward COVID-19. This is among one of the first attempts to investigate determinants of knowledge and behaviors in the public related to COVID-19 in the United States.

The general hypothesis guiding this research is that lower levels of knowledge about the coronavirus pandemic are associated with behaviors that are contrary to current guidelines that suggest that panic buying is not necessary, to avoid large gatherings, and to avoid using medical masks. Further, there are differences in knowledge and behaviors in different age groups, by sex, education level, race, income, and political party identification.

## Methods

### Participants

This cross-sectional study recruited respondents from Amazon Mechanical Turk (MTurk). MTurk is an online platform for recruiting remote workers to complete small tasks for small amounts of money. Some studies report that MTurk sample demographics are closer to the U.S. general public than typical university samples,^15,16^ and tend to be more diverse than other Internet samples^17^. MTurk provides a quick, inexpensive method to collect data from a wide cross-section of the general public.

The MTurk interface allows Requestors (myself) to advertise Human Intelligence Tasks (HITs – the survey in this case) to Workers (survey participants). I advertised for workers age 18 and older who resided in the U.S. and offered to pay them $1.00 to complete the survey. The Institutional Review Board at Michigan State University determined that this research was exempt from full board review. Participants provided consent by answering a yes-no question at the start of the survey before they could move to the first question.

### Survey

The survey was administered in two parts. The first part asked participants basic demographic characteristics including year of birth, which was used to determine age and generational membership, i.e. Baby Boomers, Gen X,^18^ education, sex, income, race, political party affiliation, and place of residence (U.S. State). Age was included to determine differences in knowledge and behavioral patterns based on age. Some reports in the U.S. essentially call-out different age groups for ignoring public health recommendations^7,19^. In addition, there are well described patterns of health literacy based on education level,^20^ and race^21^ which may not be present in a homogeneous society such as China. Political party identification is associated with many attitudes and behaviors in the U.S. related to science and science-based recommendations^22,23^. Leaders from both major parties in the U.S. have reacted differently to the COVID-19 pandemic, likely influencing those who follow them^24,25^.

The second part of the survey included 12 questions that were adapted from Zhong et al^14^ to measure knowledge about COVID-19, including clinical characteristics, transmission, and prevention and control. The knowledge questions were scored with one point for each correct question and an aggregate score calculated (range 0 to 12), with higher scores indicating more knowledge about COVID-19. Three additional questions were asked to determine participation in specific behaviors related to recommendations from CDC and/or NIH including whether participants had spent more money than usual in the last two weeks on cleaning supplies, personal hygiene products, and food (a proxy measure of hoarding), whether they had gone to any place in the last five days where there were more than 50 people present (contradicting CDC recommendations to avoid such gatherings), and if they had worn a mask when leaving the home in the last 5 days (contradicting CDC, NIH, and healthcare official guidance).

### Statistical Analyses

Sample characteristics were generated using frequency analysis and other descriptive statistics as appropriate (Table 1). Knowledge scores were compared using independent sample t-test for differences in mean score between males and females, as well as groups based on whether people had engaged in hoarding activity or not, had attended large gatherings or not, and had worn masks or not. In addition, independent sample t-tests were used to determine differences in mean age between people who had engaged in these activities or not. Analysis of Variance (ANOVA) was used to determine differences in mean knowledge scores among groups based on education, race, income, political party, and generational age groups (i.e. Baby Boomers, Gen X, etc.) (Table 2). Multivariable linear regression was used to determine which demographic characteristics influence knowledge scores, while binomial logistic regression was used to determine which characteristics influence participation in hoarding behavior, attending large group events, and using masks (Table 3). All analyses were conducted using SPSS (v.25)^26^.

**Table 1:** Participant Demographics and COVID-19 Knowledge and Behaviors.

|  |  |
| --- | --- |
| **Demographics** | **n (%)** |
| Age - mean, SD | 37.11, 11.22 |
| **Age Categories** |  |
| Baby Boomers (born 1946-1964 ) | 104 (10.1) |
| Gen X (born 1965-1976) | 140 (13.5) |
| Millenials (born 1977-1995) | 717 (69.3) |
| Gen Z (born 1996+) | 73 (7.1) |
| **Education** |  |
| High School/GED | 102 (9.9) |
| Some College | 295 (28.5) |
| Bachelor's Degree | 469 (45.4) |
| Graduate/Professional Degree | 168 (16.2) |
| **Race** |  |
| White | 784 (75.8) |
| Black/African American | 145 (14.0) |
| Asian/Pacific Islander | 69 (6.7) |
| Other | 36 (3.5) |
| **Male Sex** | 602 (58.2) |
| **Income** |  |
| \$0-\$29,999 | 232 (22.4) |
| \$30,000-\$59,999 | 366 (35.4) |
| \$60,000-\$89,999 | 2354 (22.7) |
| \$90,000+ | 201 (19.4) |
| **Political Party** |  |
| Republican | 289 (27.9) |
| Democrat | 487 (47.1) |
| Independent | 258 (25.0) |
| **Behaviors** |  |
| In the last two weeks participant reported spending more money at a grocery store/club store on cleaning supplies, personal hygiene products, or food than normal | 649 (62.8) |
| In the last five days participant reported going to any place with more than 50 people in attendance at the same time | 320 (30.9) |
| In the last five days participant reported wearing a mask when leaving home | 244 (23.6) |
| **Knowledge Questions (mean correct: 972/12 - 80%)** | **n (% Correct)** |
| The main clinical symptoms of COVID-19 are fever, fatigue, and dry cough | 948 (91.7) |
| Unlike the common cold, stuffy nose, runny nose, and sneezing are less common in persons infected with COVID-19 | 668 (64.6) |
| There currently is no effective cure for COVID-19, but early symptomatic and supportive treatment can help most patients recover from the infection | 942 (91.1) |
| Not all persons with COVID-19 will develop severe cases. Those who are elderly and have chronic illnesses are more likely to be severe cases | 886 (85.7) |
| Eating or contacting wild animals would result in infection by the COVID-19 virus | 541 (52.3) |
| Persons with COVID-19 cannot transmit the virus to others when a fever is not present | 820 (79.3) |
| The COVID-19 virus spreads via respiratory droplets of infected individuals | 917 (88.7) |
| Ordinary residents can wear general medical masks to prevent infection by the COVID-19 virus | 567 (54.8) |
| It is not necessary for children/young adults to take measures to prevent infection with COVID-19 | 878 (84.9) |
| To prevent infection with COVID-19, individuals should avoid going to crowded places and avoid public transportation | 973 (94.1) |
| Isolation and treatment of people who are infected with COVID-19 are effective ways to reduce the spread of the virus | 957 (92.6) |
| People who have contact with someone infected with the COVID-19 virus should be immediately isolated. In general, the observation period is 14 days. | 955 (92.4) |

**Table 2:**
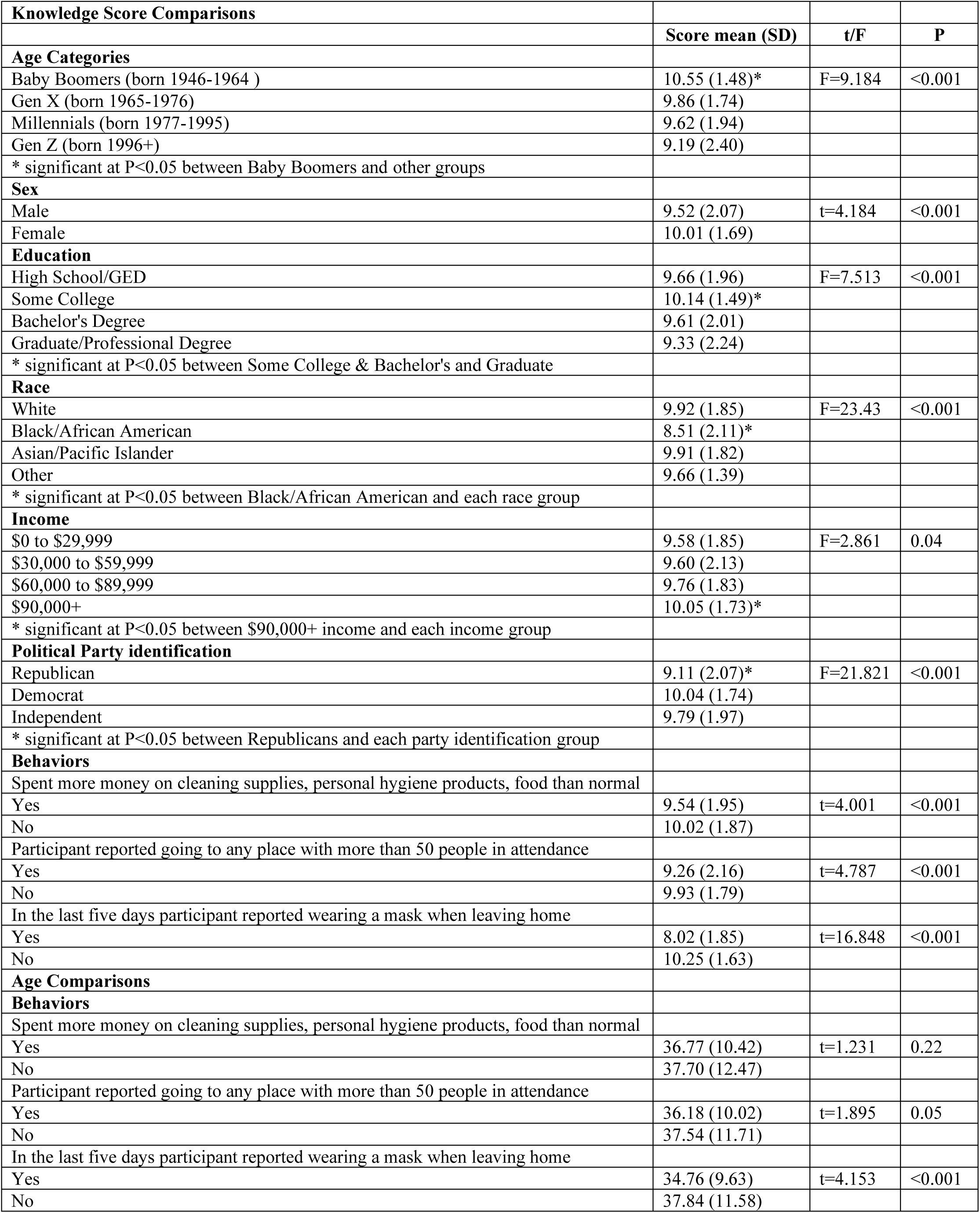
Group Comparisons of Knowledge Scores and Age.

| **Table 2: Group Comparisons of Knowledge Scores and Age** |  |  |  |
| --- | --- | --- | --- |
| **Knowledge Score Comparisons** |  |  |  |
|  | **Score mean (SD)** | **t/F** | **P** |
| **Age Categories** |  |  |  |
| Baby Boomers (born 1946-1964 ) | 10.55 (1.48)* | F=9.184 | <0.001 |
| Gen X (born 1965-1976) | 9.86 (1.74) |  |  |
| Millennials (born 1977-1995) | 9.62 (1.94) |  |  |
| Gen Z (born 1996+) | 9.19 (2.40) |  |  |
| * significant at P<0.05 between Baby Boomers and other groups |  |  |  |
| **Sex** |  |  |  |
| Male | 9.52 (2.07) | t=4.184 | <0.001 |
| Female | 10.01 (1.69) |  |  |
| **Education** |  |  |  |
| High School/GED | 9.66 (1.96) | F=7.513 | <0.001 |
| Some College | 10.14 (1.49)* |  |  |
| Bachelor's Degree | 9.61 (2.01) |  |  |
| Graduate/Professional Degree | 9.33 (2.24) |  |  |
| * significant at P<0.05 between Some College & Bachelor's and Graduate |  |  |  |
| **Race** |  |  |  |
| White | 9.92 (1.85) | F=23.43 | <0.001 |
| Black/African American | 8.51 (2.11)* |  |  |
| Asian/Pacific Islander | 9.91 (1.82) |  |  |
| Other | 9.66 (1.39) |  |  |
| * significant at P<0.05 between Black/African American and each race group |  |  |  |
| **Income** |  |  |  |
| \$0 to \$29,999 | 9.58 (1.85) | F=2.861 | 0.04 |
| \$30,000 to \$59,999 | 9.60 (2.13) | | |
| \$60,000 to \$89,999 | 9.76 (1.83) | | |
| \$90,000+ | 10.05 (1.73)* | | |
| * significant at P<0.05 between \$90,000+ income and each income group | | | |
| **Political Party identification** |  |  |  |
| Republican | 9.11 (2.07)* | F=21.821 | <0.001 |
| Democrat | 10.04 (1.74) |  |  |
| Independent | 9.79 (1.97) |  |  |
| * significant at P<0.05 between Republicans and each party identification group |  |  |  |
| **Behaviors** |  |  |  |
| Spent more money on cleaning supplies, personal hygiene products, food than normal |  |  |  |
| Yes | 9.54 (1.95) | t=4.001 | <0.001 |
| No | 10.02 (1.87) |  |  |
| Participant reported going to any place with more than 50 people in attendance |  |  |  |
| Yes | 9.26 (2.16) | t=4.787 | <0.001 |
| No | 9.93 (1.79) |  |  |
| In the last five days participant reported wearing a mask when leaving home |  |  |  |
| Yes | 8.02 (1.85) | t=16.848 | <0.001 |
| No | 10.25 (1.63) |  |  |
| **Age Comparisons** |  |  |  |
| **Behaviors** |  |  |  |
| Spent more money on cleaning supplies, personal hygiene products, food than normal |  |  |  |
| Yes | 36.77 (10.42) | t=1.231 | 0.22 |
| No | 37.70 (12.47) |  |  |
| Participant reported going to any place with more than 50 people in attendance |  |  |  |
| Yes | 36.18 (10.02) | t=1.895 | 0.05 |
| No | 37.54 (11.71) |  |  |
| In the last five days participant reported wearing a mask when leaving home |  |  |  |
| Yes | 34.76 (9.63) | t=4.153 | <0.001 |
| No | 37.84 (11.58) |  |  |

**Table 3:** Determinants of Knowledge Score and Behavior Outcomes.

| **Table 3: Determinants of Knowledge Score and Behavior Outcomes** |  |  |  |  |  |
| --- | --- | --- | --- | --- | --- |
|  | **Knowledge Score** |  | **Bought More Goods** | **Gathering >50** | **Wore Mask** |
|  | b (SE) | P | Odds Ratio (95% CI) | Odds Ratio (95% CI) | Odds Ratio (95% CI) |
| **Constant - b (SE)** | 9.90 (0.29) | <0.001 | 0.69 (0.51) | -0.08 (0.53) | 2.99 (0.73) |
| **R square** | 0.149 |  | 0.08 | 0.07 | 0.45 |
| **Knowledge Score** |  |  | 0.88 (0.81-0.95) | 0.87 (0.81-0.93) | 0.56 (0.50-0.62) |
| **Age (Reference: Baby Boomers)** |  |  |  |  |  |
| Gen X (born 1965-1976) | -0.53 (0.24) | 0.02 | 1.76 (1.03-3.01) | 1.23 (0.68-2.23) | 1.28 (0.56-2.88) |
| Millenials (born 1977-1995) | -0.64 (0.19) | 0.001 | 1.56 (1.01-2.41) | 1.35 (0.82-2.22) | 1.27 (0.63-2.54) |
| Gen Z (born 1996+) | -1.28 (0.28) | <0.001 | 0.94 (0.50-1.77) | 0.96 (0.46-1.99) | 0.83 (0.28-2.42) |
| **Male Sex** | -0.31 (0.12) | 0.007 | 0.88 (0.67-1.15) | 0.96 (0.73-1.28) | 1.35 (0.92-1.96) |
| **Education (Reference: High School/GED)** |  |  |  |  |  |
| Some College | 0.36 (0.21) | 0.09 | 1.40 (0.88-2.23) | 1.62 (0.93-2.81) | 1.23 (0.51-2.95) |
| Bachelor's Degree | -0.17 (0.20) | 0.39 | 1.88 (1.19-2.97) | 1.59 (0.93-2.72) | 4.47 (2.00-9.97) |
| Graduate/Professional Degree | -0.41 (0.24) | 0.09 | 2.11 (1.22-3.65) | 1.67 (1.46-4.87) | 7.41 (3.07-17.91) |
| **Race (Reference: White)** |  |  |  |  |  |
| Black/African American | -1.19 (0.17) | <0.001 | 1.28 (0.84-1.95) | 1.16 (0.78-1.73) | 2.48 (1.52-4.07) |
| Asian/Pacific Islander | -0.01 (0.23) | 0.97 | 1.45 (0.82-2.54) | 0.93 (0.53-1.64) | 0.62 (0.27-1.42) |
| Other | -0.19 (0.31) | 0.54 | 0.80 (0.40-1.59) | 1.11 (0.53-2.33) | 1.40 (0.53-3.67) |
| **Income (Reference: \$0-\$29,999)** | | | | | |
| \$30,000 to \$59,999 | 0.26 (0.15) | 0.09 | 1.44 (1.02-2.05) | 1.13 (0.78-1.66) | 0.99 (0.59-1.65) |
| \$60,000 to \$89,999 | 0.40 (0.17) | 0.02 | 1.44 (0.97-2.14) | 1.04 (0.68-1.59) | 1.21 (0.69-2.11) |
| \$90,000+ | 0.71 (0.18) | <0.001 | 1.54 (1.01-2.36) | 1.22 (0.78-1.90) | 0.76 (0.42-1.39) |
| **Political Party identification (Reference: Republican)** |  |  |  |  |  |
| Democrat | 0.76 (0.14) | <0.001 | 1.07 (0.77-1.49) | 0.70 (0.50-0.97) | 0.52 (0.34-0.78) |
| Independent | 0.57 (0.16) | <0.001 | 0.78 (0.54-1.12) | 0.84 (0.58-1.23) | 0.34 (0.19-0.57) |

## Results

A total of 1,070 participants completed the survey. The use of VPN networks allows people from all over the world to mimic U.S. IP addresses and so I asked for the U.S. state of residence for each participant and matched those to each IP address location to determine matches. I then excluded responses from participants whose IP address location did not match their given State. Thirty-six participants were excluded for a final sample size of 1,034. The survey was offered to MTurk workers on March 17, 2020 at 4:05 PM Eastern time, and all 1,070 responses were completed by 6:13 PM Eastern time. To set the context for the setting of the study, at the time the survey was released, there were 5,704 COVID-19 cases reported in the U.S. and 195,957 worldwide. At the date of this writing (March 24, 2020) there were 46,548 cases in the U.S. and 396,249 worldwide^27^. It is likely that these numbers vastly underrepresent the actual prevalence.

On average it took four minutes to complete the survey (equivalent to $15/hour). Participants were 37.11 years old ranging from 19 to 77, 45% had completed a bachelor’s degree, 75.8% reported white race, 58.2% were male, 35% reported income between $30,000 and $59,999, and 47.1% identified as Democrats. Additional demographic information is included in Table 1.

Results for each of the COVID-19 knowledge questions are included in Table 1. Correct answers for questions ranged from 52.3% correct (Eating or contacting wild animals would result in COVID-19 infection – False) to 94.1% (Individuals should avoid large crowds to prevent infection with COVID-19 – True). The mean knowledge score was 9.72 (standard deviation=1.93, range 0-12) for an overall correct percentage of approximately 80%, lower than the 90% correct rate that Zhong at al^14^ report in their sample of Chinese citizens at approximately two months into the outbreak.

Knowledge scores were significantly different between groups based on sex, generational ages, education, race, income, and political party identification. In general, Baby Boomers, females, those with some college, and higher incomes were more knowledgeable about COVID-19, while Black/African Americans, and Republicans were less knowledgeable (Table 2).

Regarding behaviors, participants who reported spending more money in the last two weeks, or going to gatherings with more than 50 people, or wearing masks outside the home, were less knowledgeable about COVID-19 compared to participants who did not report these activities. In addition, participants who reported the above behaviors were also significantly younger, except for increased spending where there was no significant difference in age (Table 2).

Multivariable linear regression (Table 3) results suggest several important relationships. First, compared to Baby Boomers, members of Gen X (b=-0.53, P=0.02), Millennials (b=-0.64, P=0.001), and Gen Z (b=-0.31, P<0.001) had significantly lower COVID-19 knowledge scores. Exponentiating the unstandardized parameter estimate indicates that predicted mean knowledge scores for Gen X, Millennials, and Gen Z are 42%, 53%, and 73%, respectively, lower than Baby Boomers. Black/African Americans have mean knowledge scores that are 70% lower (b=-1.19, P<0.001) compared to Whites. In general, participants with higher incomes have higher knowledge scores. Finally, Democrats (b=0.76, P<0.001), and Independents (b=0.57, P<0.001) have mean knowledge scores that are 113% and 76% higher, respectively, than Republicans.

Binary logistic regression analysis (Table 3) results reveal several predictors of each behavior. Self-reports of buying more goods than usual is negatively associated with COVID-19 knowledge. For every point increase in knowledge score, the odds of reporting unusual buying behavior decreased by 12% (OR=0.88, 95% CI:0.81-0.95). In the context of generational groups, the odds of reporting purchasing behavior increased by 76% and 56% for Gen X participants (OR=1.76, 95% CI:1.03-3.01) and Millennials (OR=1.56, 95% CI:1.01-2.41), respectively, compared to Baby Boomers. Next, people with higher education were associated with increased buying behaviors. The odds of unusual purchasing behavior increased by 88% and 111% for people with bachelor’s degrees (OR=1.88, 95% CI:1.19-2.97), and Graduate/Professional degrees (OR=2.11, 95% CI:1.22-3.65), respectively, compared to those with a high school education. Finally, those with higher incomes had increased odds of unusual purchasing behavior.

Next, for every point increase in knowledge scores, the odds of attending large gatherings in the last five days decreased by 13% (OR=0.87, 95% CI:0.81-0.93). Participants with Graduate/Professional degrees had 67% greater odds (OR=1.67, 95% CI:1.46-4.87) of attending large gatherings, compared to those with a high school education. Finally, Democrats had 30% lower odds (OR=0.70, 95% CI:0.50-0.97) of attending large gatherings compared to Republicans.

Last, for every point increase in knowledge scores, the odds of wearing a mask outside the home decreased by 44% (OR=0.56, 95% CI:0.50-0.62). The largest effect of any of the analyses revealed that those with a bachelor’s degree (OR=4.47, 95% CI:2.00-9.97), and Graduate/Professional degrees (OR=7.41, 95% CI:3.07-17.91) had 347% and 641% increased odds of wearing masks outside the home, compared to respondents with a high school education. Black/African American participants had 148% increased odds (OR=2.48, 95% CI:1.52-4.07) of wearing masks outside the home compared to Whites. Finally, Democrats (OR=0.52, 95% CI: 0.34-0.78) and Independents (OR=0.34, 95%CI:0.19-0.57) had 48% and 66% lower odds, respectively, of reporting wearing masks, compared to Republicans.

## Discussion

The PUS literature posits that an increase in knowledge leads people to understand science, and trust in the institution of science. Extending this to the current COVID-19 pandemic, I hypothesized that increased knowledge should lead to willingness to follow public health recommendations. In this sample, lower knowledge is associated with self-reports of engaging in purchasing more goods than necessary, attending gatherings of more than 50 people, and wearing medical masks outside the house. In addition, there were differences in knowledge about COVID-19 based on age group. In fact, contrary to recent U.S. media, Baby Boomers in this sample are more knowledgeable about COVID-19 than all other age groups and are less likely to engage in purchasing behavior that could be considered hoarding. In general, people who do not engage in these behaviors had significantly higher knowledge scores. Finally, people who reported attending large gatherings and wearing masks in public were younger on average.

The average knowledge score for this entire sample is about 9.72 out of 12 total points (approximately 80%) at a time about eight weeks after the first case was diagnosed in the U.S. Approximately eight weeks after the first diagnosis in China, the mean knowledge score for a sample of Chinese citizens was 10.8/12 (approximately 90%)^14^ and it was suggested that the knowledge of Chinese citizens is high because of their experiences with the SARS outbreak in the early 2000s, and the observation that this sample was relatively affluent and highly educated.

In this sample nearly 30% of people reported attending gatherings or going to places with more than 50 people in the previous five days, contrary to advice from the CDC since March 12, 2020 (survey conducted on March 17, 2020). In China, only 3.6% of people reported going to crowded places in the previous two weeks^14^. It is possible that the coordinated effort from, as well as the unchecked authority by, the Chinese government, to lockdown provinces provided most of the motivation for Chinese citizens to obey these mandates. To date, there has not been a coordinated effort by the U.S. government to lockdown the nation. There is some debate whether the federal government even the Constitutional authority and so individual states are left to make decisions about “shelter at home” policies and similar efforts. As of this writing, California, Illinois, New York, Washington, Michigan, Massachusetts, Indiana, Oregon, and West Virginia have issued stay-at-home orders. With about 1 in 3 U.S. citizens ordered to stay home it is likely in the coming weeks that fewer people will report attending large gatherings.

Use of masks is an evolving, and cultural phenomenon. In Asia, people are encouraged, and even mandated to wear masks outside the house. In China, only 2.0% of people reported not wearing masks outside the home^14^. In this sample, approximately 76% of people did not wear masks outside the home in the last five days, which is perhaps reflective of the CDC and NIH recommendations that the general public not use masks so that they are saved for front-line healthcare workers^28^. However, it is probably more likely that masks could not be found in the U.S. because of lack of supply combined with hoarding behavior^10^. Still, 24% of people reported using masks, indicating that a large section of the U.S. public chose to ignore recommendations. It is important to note that the debate on masks has changed even since this survey was conducted only six days ago. The FDA and CDC are currently debating the merits of wearing masks in public because of the understanding that many people with mild symptoms may not even know they are infected with COVID-19. Mask use could prevent infecting others by asymptomatic carriers.

Finally, political party identification significantly influenced knowledge about COVID-19 as well as behaviors related to attending large gatherings and wearing medical masks. To summarize, Republicans had lower knowledge and had higher odds of attending large gatherings and wearing masks in public compared to Democrats and/or Independents. These behaviors directly contradict recommendations by both the CDC and NIH. In the U.S. there is a widening gap in trust in science and science-based recommendations based on political party^22^ which may contribute to the observation here that Republicans are more likely to ignore recommendations about the COVID-19 response. In addition, the results reported here agree findings that there are political divisions over the role of scientific experts in policy matters^23^. That is, Democrats want expert involvement and believe scientists should be involved in policy recommendations. Conversely, Republicans believe scientists should stay out of policy debates. These attitudes may be reflected in these results that Republicans have lower knowledge about COVID-19 and have higher odds of participating in behaviors that are not recommended by authorities to stem the tide of the current pandemic.

There are some limitations to this research. First, knowledge questions are not validated and scientific knowledge is currently a moving target. For example, while the current consensus is that eating wild animals will not transmit the disease, living and working in close proximity to animals clearly influenced this outbreak and could influence future outbreaks. As such, the argument for banning wet markets in China is gaining momentum, but knowledge about proximity to animals, as opposed to using them as a food source, might be conflated. In addition, knowledge regarding who is most at risk for COVID-19 may change as the pandemic proceeds, as well as with experiences in different countries. For instance, fewer younger people in China were infected, while in the U.S. a different pattern appears to be emerging^29^. Next, this was a convenience sample of U.S. residents from every state in the country, but people were able to self-select based on their interest and experience with the topic. It is possible that sample demographics may not completely represent the U.S. public. Finally, although the survey questions were not able to be validated given the fast-moving nature of the pandemic response in the U.S., the questions do have face value in the context of the situation at the time the survey was conducted.

## Conclusions

This survey is one of the first attempts to describe determinants of U.S. public knowledge and behavioral response to the emerging COVID-19 pandemic in the United States. While knowledge about COVID-19 is generally high, there are differences in knowledge based on age, sex, education, income, race, and political party identification. These differences appear to prevent a coordinated effort at slowing the spread of the pandemic in the U.S. in these early days. Ignoring official recommendations for crowd avoidance, use of medical supplies, and purchasing behaviors that signal hoarding of goods, does not bode well for efforts to contain the spread of the virus and limit exposures to vulnerable populations. Without a coordinated national response, it is likely the U.S. will experience a longer, more drawn out battle than if such coordination would occur.

## Data Availability

Original data can be requested from the author.

## Acknowledgements

This study was funded by faculty development funds from the Master of Public Health Program, Division of Public Health, College of Human Medicine, Michigan State University.

The author reports no conflicts of interest.

## Reference List

1. World Health Organization. Novel coronavirus (2019-nCoV) situation report - 1. https://www.who.int/docs/default-source/coronaviruse/situation-reports/20200121-sitrep-1-2019-ncov.pdf?sfvrsn=20a99c10_4. Published January 20, 2020. Accessed March 24, 2020.

2. BBC. Lockdowns rise as China tries to control virus. https://www.bbc.com/news/world-asia-china-51217455. Published January 23, 2020. Accessed march 24, 2020

3. Washington State Department of Health. Novel coronavirus outbreak 2020. Updated March 3, 2020. https://www.doh.wa.gov/emergencies/coronavirus. Published March 3, 2020. Accessed March 24, 2020.

4. The New York Times. Coronavirus may have Spread in U.S. for Weeks, Gene Sequencing Suggests. https://www.nytimes.com/2020/03/01/health/coronavirus-washington-spread.html. Published March 1, 2020. Accessed March 24, 2020

5. Morrison C. “Trump declares coronavirus outbreak a public emergency, will ban foreign travel from China and quarantine US citizens”. Washington Examiner. https://www.washingtonexaminer.com/news/trump-administration-declares-coronavirus-outbreak-a-public-health-emergency. Published January 31, 2020. Accessed March 24, 2020.

6. Centers for Disease Control and Prevention. How to Protect Yourself. https://www.cdc.gov/coronavirus/2019-ncov/prepare/prevention.html. Accessed March 24, 2020

7. Young A and Blondin A. What coronavirus? College students flood Myrtle Beach to party as COVID-19 spreads. The Charlotte Observer. https://www.charlotteobserver.com/news/coronavirus/article241350261.html. Published March 19, 2020. Accessed March 24, 2020.

8. Thorbecke C. Americans hoarding hand sanitizer, face masks and oat milk amid coronavirus fears. https://abcnews.go.com/Business/americans-hoarding-hand-sanitizer-face-masks-amid-coronavirus/story?id=69385946. Published March 4, 2020. Accessed march 24, 2020.

9. Brooks B and Hay A. Hoarding in the USA? Coronavirus sparks consumer concerns. https://www.reuters.com/article/us-china-health-usa-hoarding-idUSKCN20M37V. Reuters. Published February 28, 2020. Accessed March 24, 2020.

10. Yen H and Madhani A. Trump calls on Americans to cease hoarding food, supplies. https://apnews.com/c94f009b46741786a0fa3307a4841ba7. Associated Press. Published March 15, 2020. Accessed March 24, 2020.

11. Solomon J. Reception and rejection of scientific knowledge: choice, style and home culture Public Understanding of Science. 1993;2:111–121.

12. Allum N, Sturgis P, Tabourazi D, and Brunton-Smith I. Science knowledge and attitudes across cultures: a meta-analysis. Public Understanding of Science. 2008;17:35–54.

13. Miller JD. Public understanding of, and attitudes toward, scientific research: what we know and what we need to know. Public Understanding of Science. 2004;13:273–294.

14. Zhong B, Luo W, Zhang QQ, Liu XG, Li WT, Li Yi. Knowledge, attitudes, and practices toward COVID-19 among Chinese residents during rapid rise period of COVID-19 outbreak: a quick online cross-sectional survey. International Journal of Biological Sciences. 2020;16(10):1745–1752.

15. Mason W and Suri S. Conducting behavioral research on Amazon’s Mechanical Turk. Behavior Research Methods. 2012;44(1):1–23.

16. Paolacci G, Chandler J and Ipeirotis PG. Running experiments on Amazon Mechanical Turk.” Judgment and Decision Making. 2010;5(5):411–419.

17. Buhrmester M, Kwang T, and Gosling SD. “Amazon’s Mechanical Turk: A new source of inexpensive, yet high-quality, data? Perspectives on Psychological Science. 2011;6(1) 3–5.

18. Center for Generational Kinetics. Generations. https://genhq.com/FAQ-info-about-generations/. Accessed March 18, 2020.

19. Schulman M. Convincing Boomer parents to take the coronavirus seriously. https://www.newyorker.com/culture/culture-desk/convincing-boomer-parents-to-take-the-coronavirus-seriously. The New Yorker. Published March 16, 2020. Accessed March 24, 2020.

20. Sun X, Shi Y, Zeng Q, et al. Determinants of health literacy and health behavior regarding infectious respiratory diseases: a pathway model. BMC Public Health. 2013;13(261): doi.org/10.1186/1471-2458-13-261.

21. Chaudhry SI, Herrin J, Phillips C, et al. Racial disparities in health literacy and access to care among patients with heart failure. Journal of cardiac failure, 2011;17(2):122–7. Doi:10.1016/j.cardfail.2010.09.016.

22. Krause NM, Brossard D, Scheufele DA, Xenos MA, Franke K. Trends—Americans’ trust in science and scientists, Public Opinion Quarterly. 2019;83(4):817–36. https://doi.org/10.1093/poq/nfz041.

23. Funk C, Hefferon M, Kennedy B, and Johnson C; Pew research Center. Trust and mistrust in Americans’ views of scientific experts. https://www.newswise.com/pdf_docs/156460235934074_Embargoed%20REPORT%20trust%20scientists%207-30-19.pdf. Published August 2, 2019. Accessed March 24, 2020.

24. Abramowitz AI and Webster SW. Negative partisanship: Why Americans dislike parties but behave like Rabid Partisans. Political Psychology. 2018;39:119–135. https://doi.org/10.1111/pops.12479.

25. Mason L and Wronski J. One tribe to bind them all: how our social group attachments strengthen partisanship. Political Psychology. 2018;39:257–277. doi:10.1111/pops.12485.

26. IBM Corp. Released 2017. IBM SPSS Statistics for Windows, Version 25.0. Armonk, NY: IBM Corp.

27. Johns Hopkins University. Coronavirus COVID-19 Global Cases by the Center for Systems Science and Engineering (CSSE) at Johns Hopkins University (JHU). https://gisanddata.maps.arcgis.com/apps/opsdashboard/index.html#/bda7594740fd40299423467b48e9ecf6. Published daily. Accessed on March 17, 18, and 23, 2020.

28. U.S. Food and drug Administration. Respirators and surgical masks. https://www.fda.gov/medical-devices/personal-protective-equipment-infection-control/n95-respirators-and-surgical-masks-face-masksN95. Accessed March 24, 2020.

29. Cha AE. Younger adults are large percentage of coronavirus hospitalizations in United States, according to new CDC data. https://www.washingtonpost.com/health/2020/03/19/younger-adults-are-large-percentage-coronavirus-hospitalizations-united-states-according-new-cdc-data/. The Washington Post. Published March 19, 2020. Accessed March 24, 2020.

